# COVID-19 in Londrina-PR-Brazil: SEIR Model with Parameter Optimization

**DOI:** 10.1101/2021.07.27.21261227

**Authors:** Eliandro R. Cirilo, Paulo L. Natti, Neyva M. L. Romeiro, Pedro H. V. Godoi, Andina A. Lerma, Vitor P. Matias

**Affiliations:** Depto. de Matemática, UEL, Londrina, Paraná, Brazil

**Keywords:** COVID-19, Londrina-PR, SEIR Model, Parameter Optimization, ODE, Bio-Mathematics

## Abstract

The first cases of COVID-19 in Londrina-PR were manifested in March 2020 and the disease lasts until the present moment. We aim to inform citizens in a scientific way about how the disease spreads. The present work seeks to describe the behavior of the disease over time. We started from a compartmental model of ordinary differential equations like SEIR to find relevant information such as: transmission rates and prediction of the peak of infected people. We used the data released by city hall of Londrina to carry out simulations in periods of 14 days, applying a parameter optimization technique to obtain results with the greatest possible credibility.

## Introduction

The pandemic caused by COVID-19, a disease caused by the SARS-CoV-2 virus, has been responsible for a major health crisis worldwide. It is already in the public domain that transmission is potentially through contact, with confirmed cases on all continents (BBC NEWS, 2021). In this context, COVID-19 exposed public health policies to a new challenge.

In Brazil, the first case of COVID-19 was registered on February 26, 2020. Until October 23 of the same year, less than eight months after the first case, Brazil had already registered more than five million cases, surpassing one hundred and fifty thousand deaths, according to data from the Ministry of Health (BRASIL, 2020). This represents around 13% of confirmed cases worldwide, and close to 14% of deaths in the world, if we compare with data from the World Health Organization (WHO, 2020a).

During this period of exponential growth in the number of cases, governments have increasingly searched for scientific indicators that can be used to understand the dynamics of the spread of the disease, at various scales, and for making critical executive decisions that generate great impacts on society. For this, multidisciplinary teams have been formed within the government to generate studies and support the decision-making process with scientific data.

In addition to the teams formed by projects linked to the government, the scientific community has been mobilized to prepare studies, in different areas of knowledge, to help in the understanding of COVID-19 and its impacts on society.

This work aims to inform the population, based on COVID-19 data observed in the city of Londrina-PR, from the perspective of the SEIR model (BRAUER;CASTILLO-CHAVEZ, 2012).

### COVID-19

The first records of the SARS-CoV-2 virus come from Wuhan, China, where an outbreak of an unknown disease with symptoms associated with pneumonia was observed. On January 4, 2020, the World Health Organization reported the case on its social media, announcing that it would investigate the disease (WHO, 2020f). On Jan. 12, Chinese authorities announced the discovery of the disease-causing virus, with information confirmed by the World Health Organization (WHO, 2020b). The names of the disease (COVID-19) and the virus (SARS-CoV-2) were officially assigned on February 11 (WHO, 2020e). On March 11, the World Health Organization characterizes the disease as a pandemic (WHO, 2020c).

According to the World Health Organization, the most common symptoms of COVID-19 are fever, dry cough and fatigue. Less common symptoms include headaches and loss of smell. For 80% of those infected, recovery took place without the need for hospital treatment, while for the remaining 20%, a more intense intervention is necessary. Although comorbidities such as high blood pressure, lung problems or cancer are linked to a more serious development of the disease, anyone, regardless of age or medical history, can develop severe symptoms (WHO, 2020d).

In the absence of a specific medication or vaccine that is widely available for COVID-19, prevention is still the most effective method of preventing the disease. Regarding preventive methods, social isolation, the use of masks and regular hand hygiene are the most effective to mitigate the spread of the disease.

### Londrina

The city of Londrina, located in the state of Paraná, in Brazil, is the second most populous city in the state, with an estimated population in 2019 of 569,733 inhabitants (IBGE, 2020a), only smaller than the population of Curitiba (state’s capital). Social interaction between people is a feature of the city. Services are the second largest source of GDP in Londrina, while the agro-industrial sector is the main source (IBGE, 2020b). In addition, Londrina is an important education center in Brazil, with the presence of federal, state and private universities. In this way, social isolation measures have great impact on the daily life of Londrina society.

The first case of COVID-19 in Londrina was registered on March 17, 2020 (RADIO PAIQUERE, 2020). On the same day, the Public Health Emergency Operations Center (COESP) was established, with representatives from each hospital in the city, to manage the actions related to the pandemic in the city (LONDRINA, 2020b). From March 31, the Londrina Health Department (LHD) started to release a daily bulletin^1^ with the situation of the cases of COVID-19 in the city. Until October 23, the city had 12,056 confirmed cases, with 301 deaths (LONDRINA, 2020a).

Thus, with the current increase in the frequency of new cases of COVID-19 in Londrina, this paper aims to contribute and present a quantitative analysis of the disease in this city. In this context, the SEIR mathematical epidemiological model (BRAUER;CASTILLO-CHAVEZ, 2012) was used to show estimates of the behavior of the disease, taking as input data for the model those published daily by the Londrina Health Department (LHD) at the address <https://saude.londrina.pr.gov.br/index.php/dados-epidemiologicos/boletim-informativo.html>.

### Materials and Methods

In this chapter we introduce the SEIR epidemic model and the considerations taken for its application to the COVID-19 problem in Londrina. Such models were introduced by W.O. Kermack and A.G. McKendrick between 1927 and 1933, and have been used and extended to the present day.

### SEIR Model

The SEIR model is an epidemic compartment model, that is, the total population considered in the study is divided into subpopulations (compartments), according to the state of interaction of its individuals with the disease. The transition (dynamics) of individuals between these compartments can be described by a set of ordinary differential equations.

The SEIR model has four compartments that divide the population into subpopulations called: Susceptible (S), Exposed (E), Infected (I) and Removed (R), as illustrated in Figure1.

**Figura 1.**
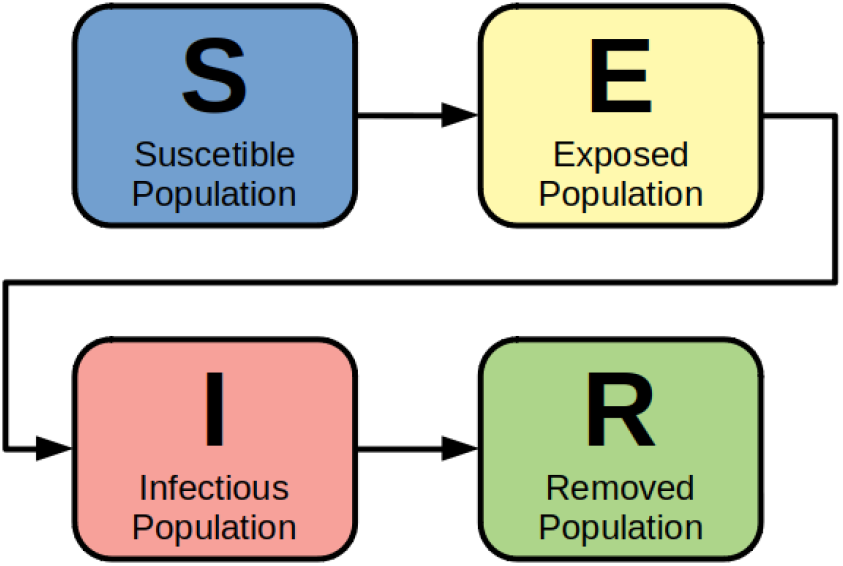
SEIR model compartments. **Source:** The authors.

In compartment (S) are healthy individuals who do not have immunity to the disease-causing SARS-CoV-2 virus. Individuals (S) can move to compartment (E) if they have contact with the virus, possibly due to random contacts with infected individuals.

In compartment (E) we have individuals infected with the virus, but still in the initial stage of the disease. They do not transmit the virus to others, since they are in the incubation period.

Compartment (I) contains infected individuals who experience a more advanced stage of the disease. They are able to transmit the virus to other individuals. In this compartment, the main symptoms associated with Covid-19 are observed, however it may also happen that some individuals are asymptomatic. After 14 days, on average, people from compartment (I) are moved to compartment (R).

Finally, compartment (R) contains all individuals who had the disease, individuals who recovered, and those who died. By hypothesis, the model assumes that the Recovered individual acquires immunity to the virus after the disease, and therefore does not become a Susceptible individual again.

Other compartments could be added to enrich the model, however the SEIR model fits the specifics and scope of this work. Thus, the compartments (S), (I) and (R) can be compared to official public data, allowing to obtain predictions about the evolution of the epidemic, based on numerical simulations. On the other hand, although compartment (E) does not have known official data, it was maintained, since the dynamics and incubation period of the disease are well established in current studies.

We also argue that the objective of this work is to estimate the number of infected (I) over time, so we opted a priori to keep the removed compartment (R) unified, that is, without distinguishing between removal due to cure or death, considering that both situations leave the infected compartment (I).

Modeling the compartmental transitions in the form of differential equations, we obtain the following system

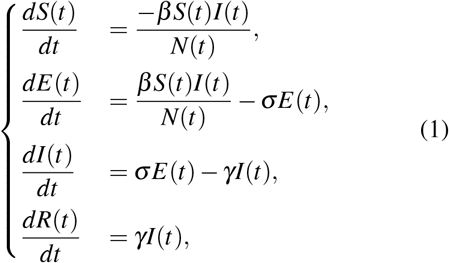

with *S*(*t*), *E*(*t*), *I*(*t*), and *R*(*t*) being the quantities of Susceptible, Exposed, Infected, and Removed individuals, over time, respectively. The quantity *N*(*t*) defines the total population over time. Finally, *β* (transmission rate), *σ* (latency/incubation rate), and *γ* (recovery rate) are parameters that need to be adjusted properly to simulate the spread of the disease in Londrina. In this work, it is considered for the average latency period *σ* ^-1^ = 5 days, while for the average recovery period *γ*^-1^ = 9 days (LAUER *et*. al., 2020).

The system of differential equations (1) will serve as the basis for our analysis of the development of COVID-19 in the city of Londrina.

From compartmental epidemic models one can deduce a value usually called the basic reproduction number, or *R*_*t*_ (BRAUER;CASTILLO-CHAVEZ, 2012), *given by*

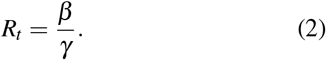

The value *R*_*t*_ can be interpreted as the average number of transmissions caused by an infected individual, before it is removed. Thus, a value *R*_*t*_ ≥ 1 indicates that each infected individual, during their infectious period, transmitted the virus to at least one individual, indicating that the epidemic is growing. On the other hand, *R*_*t*_ < 1 indicates that the epidemic is decreasing and tends to disappear.

### Modeling for Simulations

We know that the COVID-19 disease is not fully understood by the scientific community. We also observe fluctuations in the counts of the daily bulletins publish by Londrina Health Department. Therefore, in this work, we chose to carry out simulations considering periods of 14 days. With this strategy to observe the spread of the disease, simulations are more reliable, and decision-making at the government level can be more accurate

First, we measure the time *t* in days, in order to facilitate the comparison of simulation results with data from daily bulletins.

According to (IBGE, 2020a), the accumulated population growth in Londrina in the last ten years was 13.5%, so we consider, for simplicity, that the total population of the city is constant in the period of the simulations. In this way, we write that

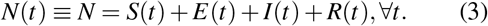

With the values of *σ* and *γ* defined (LAUER *et. al*., 2020), we need to adjust the parameter *β* (transmission rate) for the city of Londrina. We used the data provided by the Londrina Health Department (LHD) to calibrate the *β* parameter. In addition, *β, σ* and *γ* will be considered constant in each 14-day simulation period. Note that *β* and *γ* being constant in each simulation period, the value of *R*_*t*_ will also be constant in each period. The *β* optimization process will be described in the following section.

We applied the SEIR model at 14-day periods from April 27, 2020^2^. In order to apply the SEIR model, it is necessary to define the initial values of the differential equation variables for each 14-day intervals. These values are taken from the sources indicated in the Table 1. We emphasize that the subindex *I* means initial.

**Tabela 1.** Data source for initial values of SEIR model variables.

| Initial Value | Source |
| --- | --- |
| $S_I$ | $N - E_I - I_I - R_I$ |
| $E_I$ | simulated value |
| $I_I$ | LHD data |
| $R_I$ | LHD data |

**Source:** The authors.

The values of *I*_*I*_ and *R*_*I*_ were taken from the Londrina Health Department^3^. The value of *S*_*I*_ is calculated as described in Table 1, which guarantees the conservation of the equation (3), whereas the value of *E*_*I*_ is calculated by numerical simulation of the differential equation system (1).

To obtain *E*(*t*) we defined the following strategy: for the first simulation, from 04/27 to 05/10, we took *E*_*I*_ = 0, due to the low number of infected on 04/27. For the second simulation we assumed the value *E*_*I*_ as the value given by differential equation system (1) on 05/10, obtained in the first simulation. Thus, successively, the value of *E*_*I*_ of each simulation is defined by the previous simulation.

The equations were discretized via the Finite Difference Method (CUMINATO;MENEGUETTE JUNIOR,2013), and solved by the technique known as QuasiLinear (CIRILO;PETROVSKII;ROMEIRO;NATTI, 2019) through a computer application developed in GFORTRAN (GNU, 2021). Numerical modeling will follow the same process proposed in (CIRILO *et. al*., 2020), but with an optimization process for *β* as detailed below.

### β Parameter Optimization Process

The value of *β* is estimated in each simulation. Note that in the process of value optimization of the *β* parameter, only the data referring to the days in the period are used. Thus, we use the coefficient of determination *R*^2^ to compare the infected curve *I*(*t*) generated by the numerical simulation with the official data from the Municipal Health Department of Londrina, and then estimate the optimized *β* value, as described in (DEVORE, 2012).

In the simulations we observed that a correction of order 10^−4^ in *β* generated, on average, a lower order change in the value of *R*^2^. Therefore, we choose to represent *β* with three decimals.

An automated process was programmed into GFORTRAN to run the 14-day simulations. The assumed initial value for *β* was 0.001.For this value of *β*, we calculate the value of *R*^2^ by comparing the simulations with available official data. Next, we incremented the value of *β* and repeated the comparison process. The automated process ends when we obtain for *R*^2^ the closest value to 1, which means the best fit with the experimental data. In this way we find the optimized *β* value in this 14-day period. Figure 2 illustrates the optimization process.

**Figura 2.**
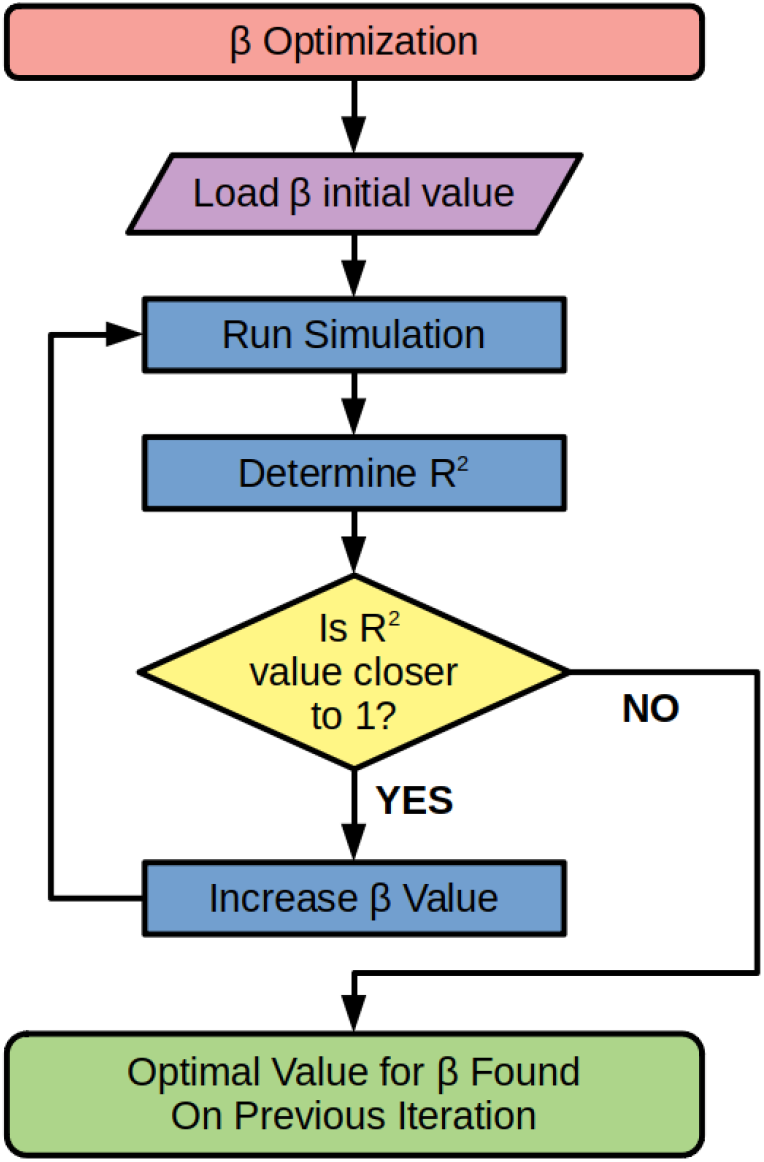
Flowchart of the *β* parameter optimization process. **Source:** The authors.

## Results

The Table 2 presents the simulation labels with their 14-day date ranges, the corresponding optimized *β* values, and the initial data with respect to exposed, infected, and removed individuals. For example, the first line refers to the first simulation (S01) of the data taken in the period of 04/27/2020 ∼ 05/10/2020, whose optimal value of *β* found was 0.184, with the number of people in the compartments (E), (I), (R), on 04/27/2020, equal to 0, 18 and 81, respectively. Similarly, we obtain the rest of the data up to simulation S13.

**Tabela 2.** Values used and obtained for the simulations.

| | Simulation | $\beta$ | $E_I$ | $I_I$ | $R_I$ |
| --- | --- | --- | --- | --- | --- |
|  | S01 04/27 ~ 05/10 | 0.184 | 0 | 18 | 81 |
|  | S02 05/10 ~ 05/23 | 0.539 | 14 | 10 | 102 |
|  | S03 05/23 ~ 06/05 | 0.114 | 143 | 113 | 142 |
|  | S04 06/05 ~ 06/18 | 0.194 | 95 | 167 | 373 |
|  | S05 06/18 ~ 07/01 | 0.076 | 214 | 229 | 700 |
|  | S06 07/01 ~ 07/14 | 0.182 | 1001 | 213 | 1133 |
|  | S07 07/14 ~ 07/27 | 0.058 | 232 | 302 | 1755 |
|  | S08 07/27 ~ 08/09 | 0.118 | 90 | 284 | 2629 |
|  | S09 08/09 ~ 08/22 | 0.175 | 143 | 276 | 3474 |
|  | S10 08/22 ~ 09/04 | 0.128 | 287 | 478 | 4192 |
|  | S11 09/04 ~ 09/17 | 0.025 | 327 | 486 | 6049 |
|  | S12 09/17 ~ 09/30 | 0.252 | 65 | 227 | 8103 |
|  | S13 09/30 ~ 10/13 | 0.001 | 381 | 304 | 9888 |

Source: The authors.

Our numerical model allows these values to be updated, so that every 14 days a new simulation can be performed and added to Table 2.

In addition, the results allow the creation of videos demonstrating the change in the curve of infected people in the city, allowing a dynamic visualization of the results, which allows a better understanding by the citizens. In the images 3 and 4 we show frames from these videos, demonstrating the shift of the *I* curve. We emphasize that the circles refer to the data released by the Health Department of Londrina, while the continuous curves result from the simulation of our model, which illustrate the past and the future from a day taken as a reference. In particular, for example on the reference date 05/23/2020, in Figure 3, the black curve before this date represents a consolidated result, which does not change anymore, while the blue curve simulates the prediction of the potential for infection.

**Figura 3.**
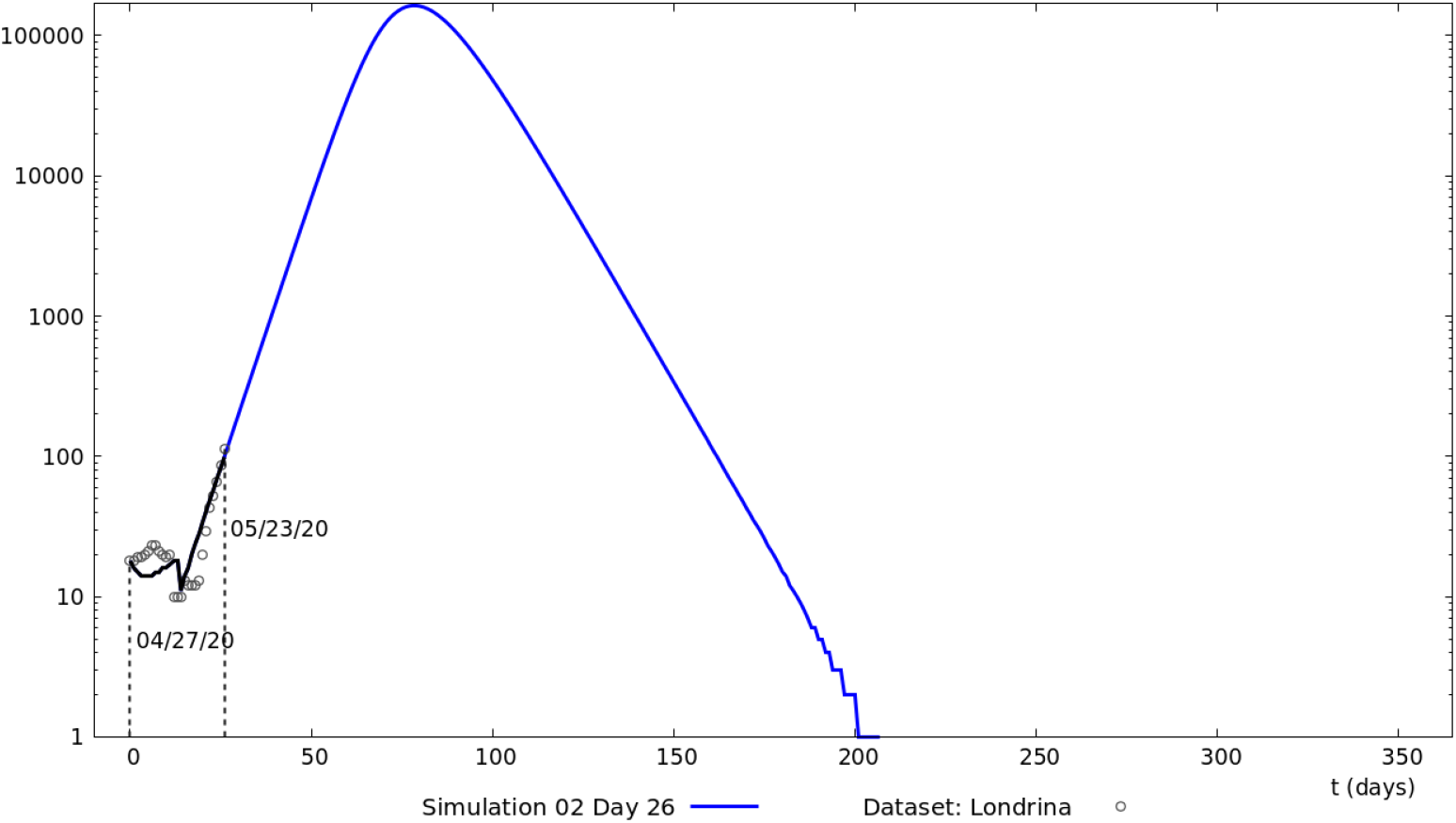
Forecast of Infected from 05/23/2020 (the 26th day of the epidemic). **Source:** The authors.

**Figura 4.**
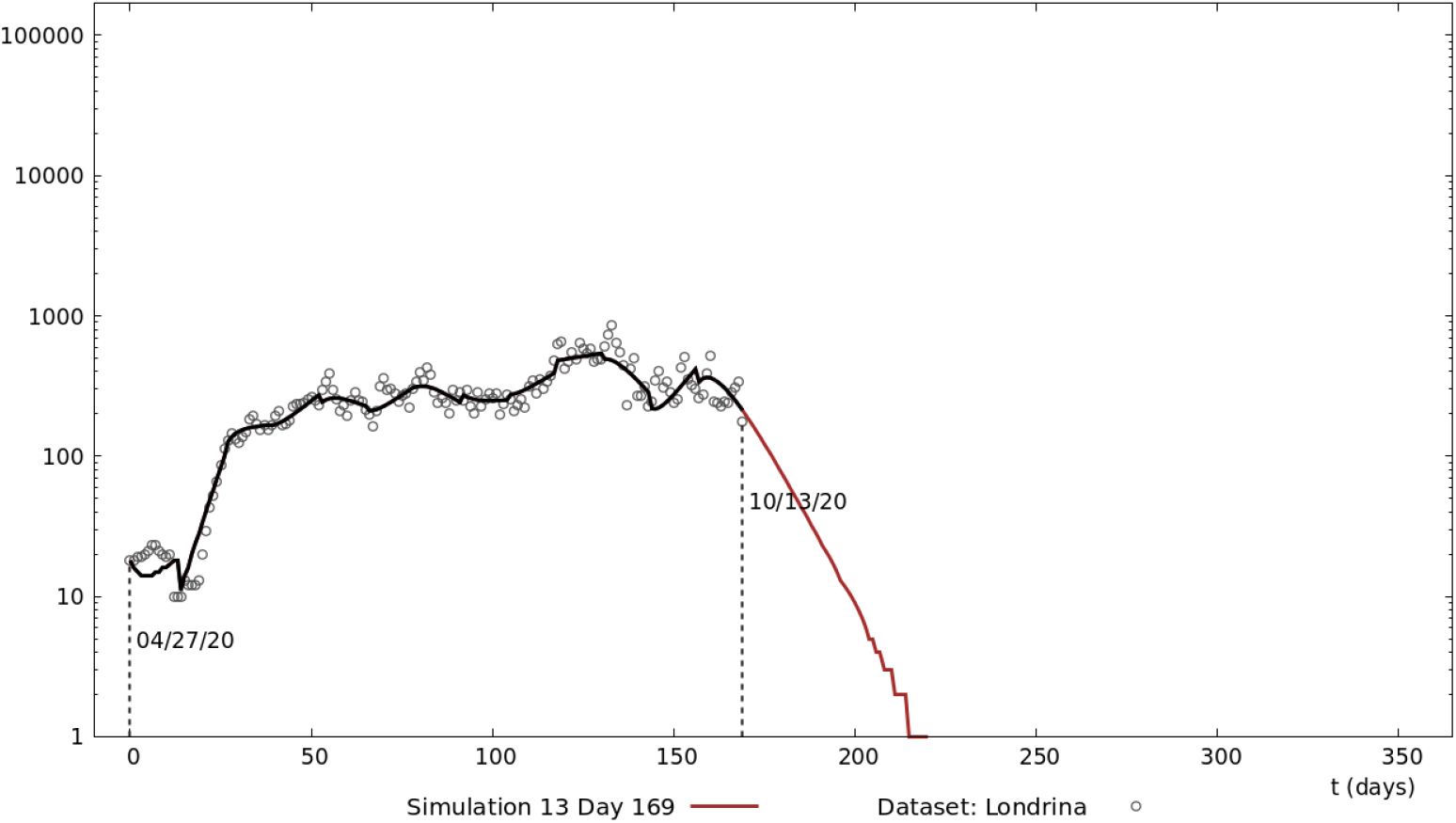
Forecast of Infected from 10/13/2020 (the 169th day of the epidemic). **Source:** The authors.

Figure 5 shows the evolution of the number of individuals infected with COVID-19. Our numerical simulations are presented, for periods of 14 days, consolidated, and the data released by the Health Department of Londrina (HDL). Note that for the first 4 periods we have a persistence of growth, and then a period of fluctuations with a slight upward trend is consolidated. This behavior is cha-racteristic of epidemics that spread, while the population reacts to contain it. Until the 195th day of the epidemic, we do not see any downward trend in the number of infected.

**Figura 5.**
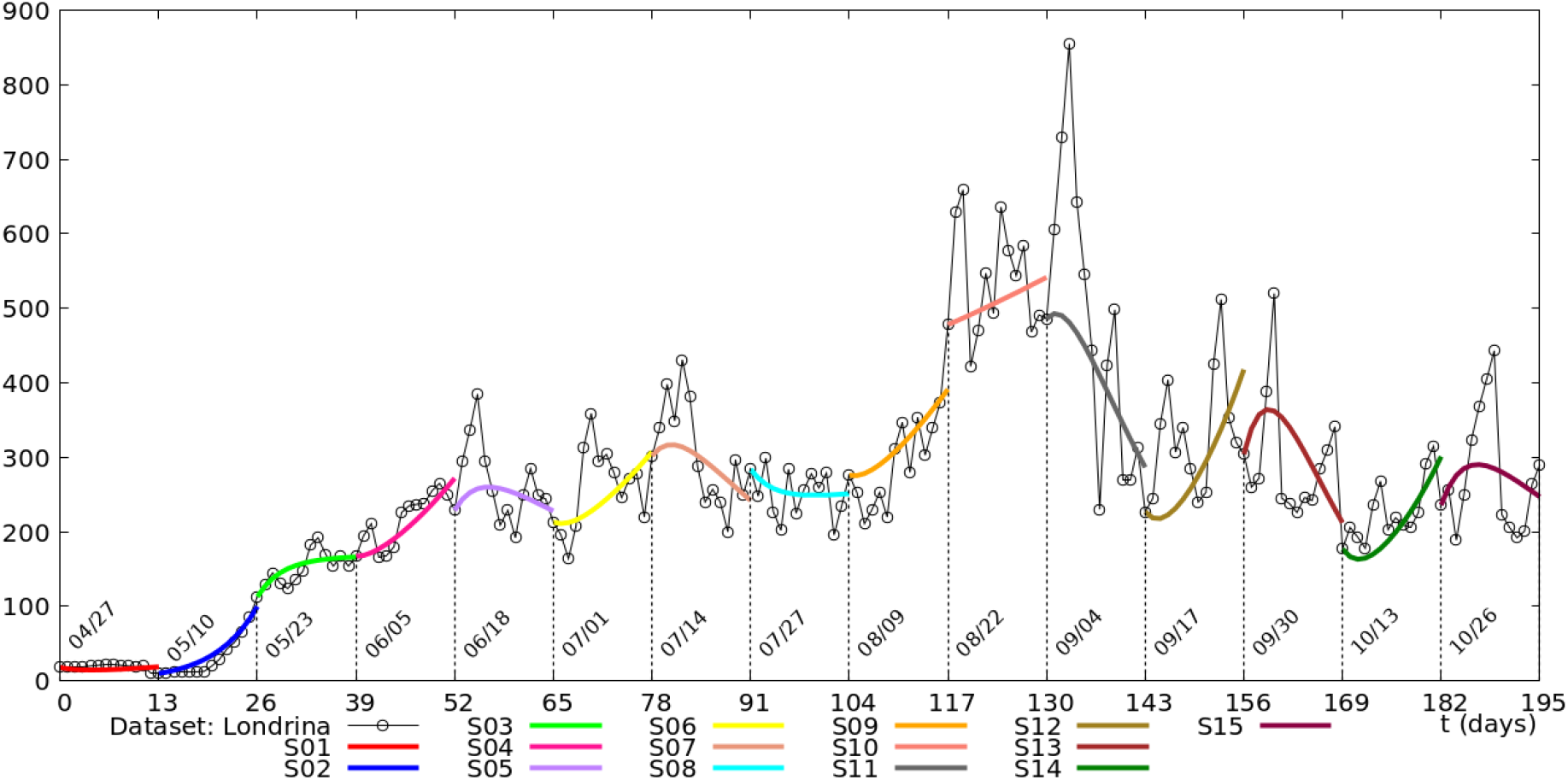
Numerical simulations, in periods of 14 days, of the number of Infected until the current moment (the 195th day of the epidemic) **Source:** The authors.

In Figure 5, simulations S01 and S02 describe an initial period of infected growth. In particular, S02 indicated an exponential growth in the number of Covid-19 cases, a common behavior in the beginning of epidemics. In the next moment, indicated by simulations S03 and S04, there was a slowdown in the growth of infected individuals. Simulations from S05 to S08 showed relative stability of the epidemic, followed by a sudden increase in the number of confirmed cases in the municipality, shown in simulations S09 and S10. This behavior does not extend to the following period, when the number of cases decreases rapidly in the S11 simulations, evidencing a strong previous intervention of HDL. This behavior does not extend to the following period, when the number of cases decreases rapidly in the S11 simulations, evidencing a strong previous intervention of HDL. Finally, we again have an oscillatory behavior indicating epidemic stability, as can be seen in simulations S12 and S15.

Figure 6 displays the same data as Figure 5, but in logarithmic scale for better visualization. Note that peaks estimated by our simulations show a tendency to soften the absolute peak values. However, unless a vaccine against COVID-19 is approved, the population will still have a long period of living with the disease.

**Figura 6.**
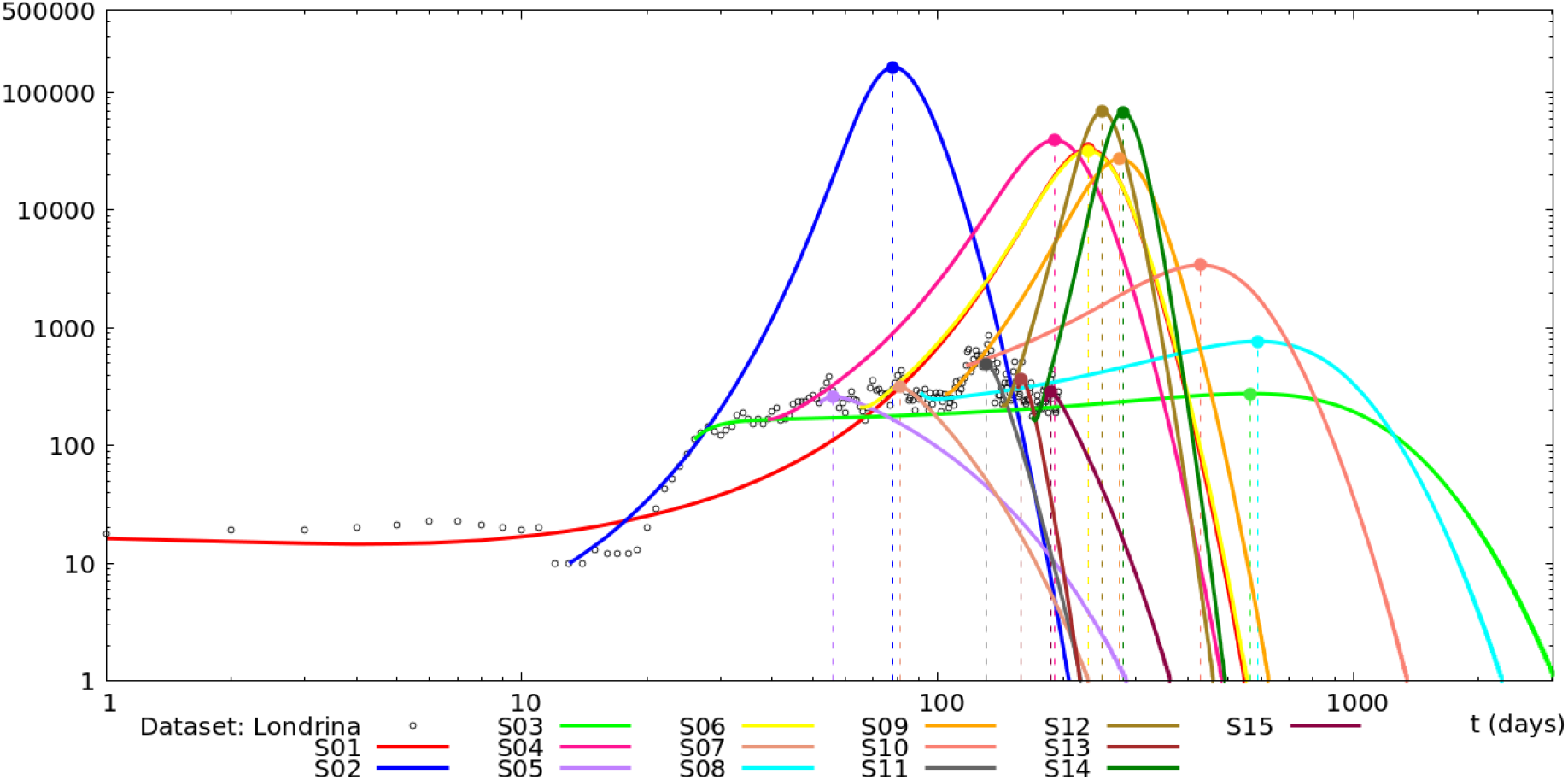
Numerical simulations on a logarithmic scale of the number of infected individuals over 14-day periods. **Source:** The authors

Table 3 presents the values of the basic reproduction number *R*_*t*_ calculated for each simulation. Note that initially we have a high value of *R*_*t*_, simulations S01 and S02 (27/04/2020 ∼ 05/23/2020), with a high rate of infection in the population. For example, for S02, we got *R*_*t*_ = 4, 851, which means that 1 infected would have the potential to infect almost 5 individuals. If this rate were maintained, this would imply affecting citizens by the disease with a peak of approximately 163,671 individuals, as can be seen in the simulated estimates in Table 3 and Figure 6. From simulations S03 to S11 (05/23/2020 ∼ 09/17/2020) the values of *R*_*t*_ oscillate relatively close to 1, indicating that the actions taken by government agencies managed to contain the spread of the disease.

**Tabela 3.**
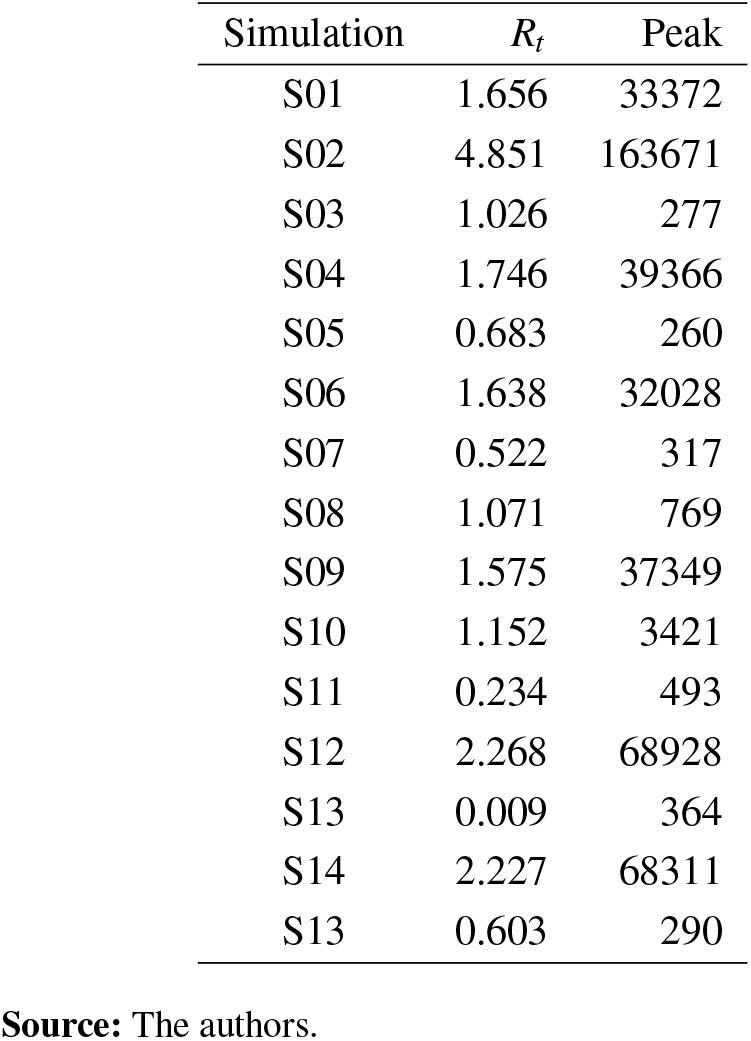
Some values obtained in numerical simulations in each 14-day period.

On the other hand, in the simulation periods S12 to S15 (09/17/2020 ∼ 11/08/2020) there are changes in the amplitude of the oscillations. Due to the long period of social contentions there is a natural relaxation, as can be seen in the social isolation indices calculated using mobile phone data (INLOCO, 2020). For example, we can observe that the spread of the disease intensifies in the simulation period S12 (09/17/2020 ∼ 09/30/2020), *R*_*t*_ = 2, 268, so that 1 infected individual would have the potential to infect approximately 2 others individuals.

Figure 7 presents the scenario as the city of Londrina-PR is reacting against COVID-19 in terms of *R*_*t*_. The black dots are estimated values for *R*_*t*_, and the continuous curve presents its trend. The violet horizontal dashed line, with a value of 1, represents the threshold for disease extinction or persistence. We emphasize that *R*_*t*_ < 1 implies having the epidemic under control. On the other hand, the maintenance of this condition is not simple, since the disease has a fast spreading capacity due to social interactions, work activities, recreational activities, among others. Furthermore, the economic maintenance of the city is important. In this context, the monitoring of *R*_*t*_ is necessary so that preventive actions can be quickly adopted by municipal authorities. In this way, the population of the city will be able to minimally experience the consequences of the disease, as long as there are no vaccines approved for COVID-19.

**Figura 7.**
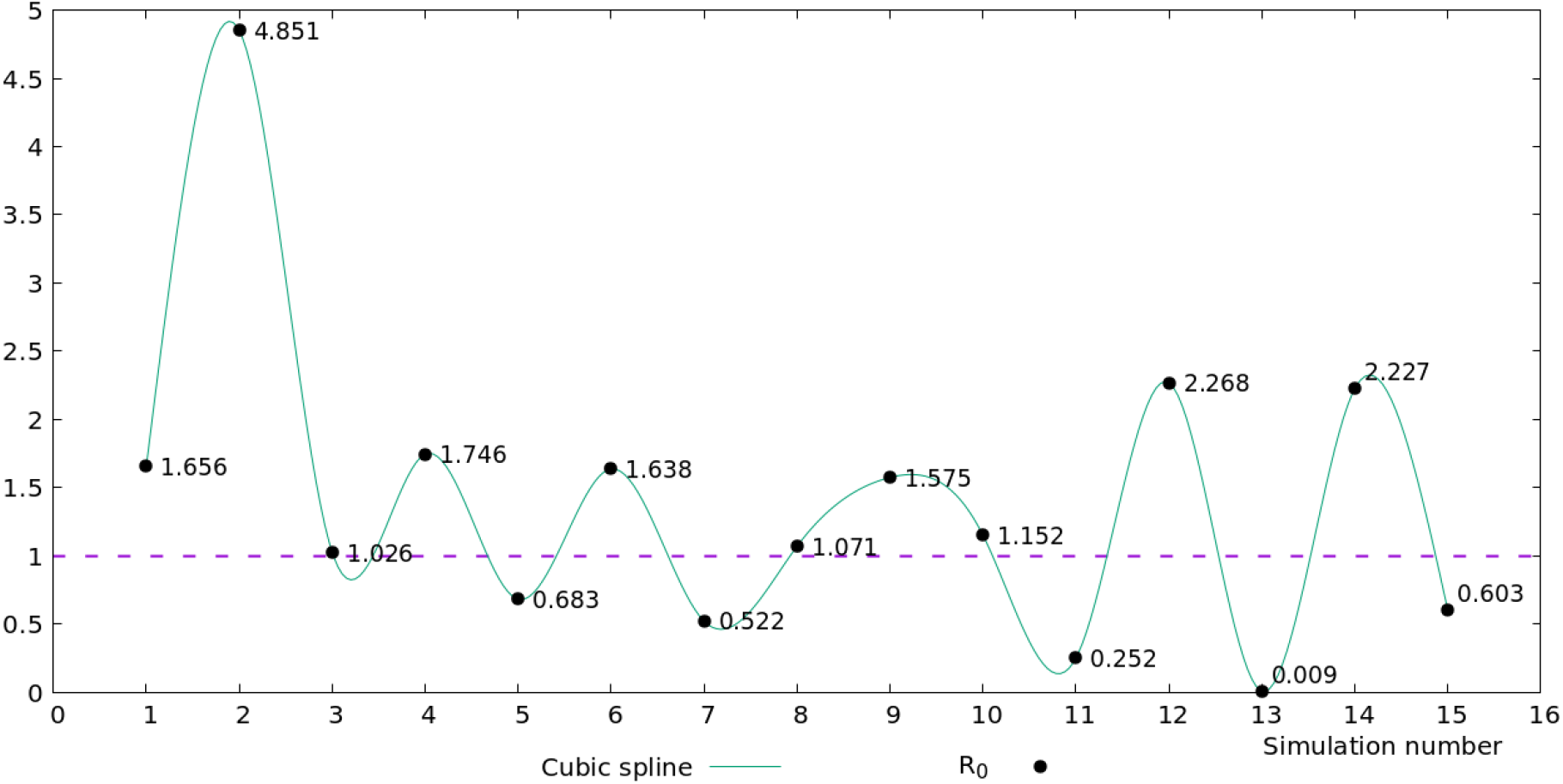
Values of *R*_*t*_ obtained in numerical simulations in each 14-day period. **Source:** The authors.

**Figura 8.**
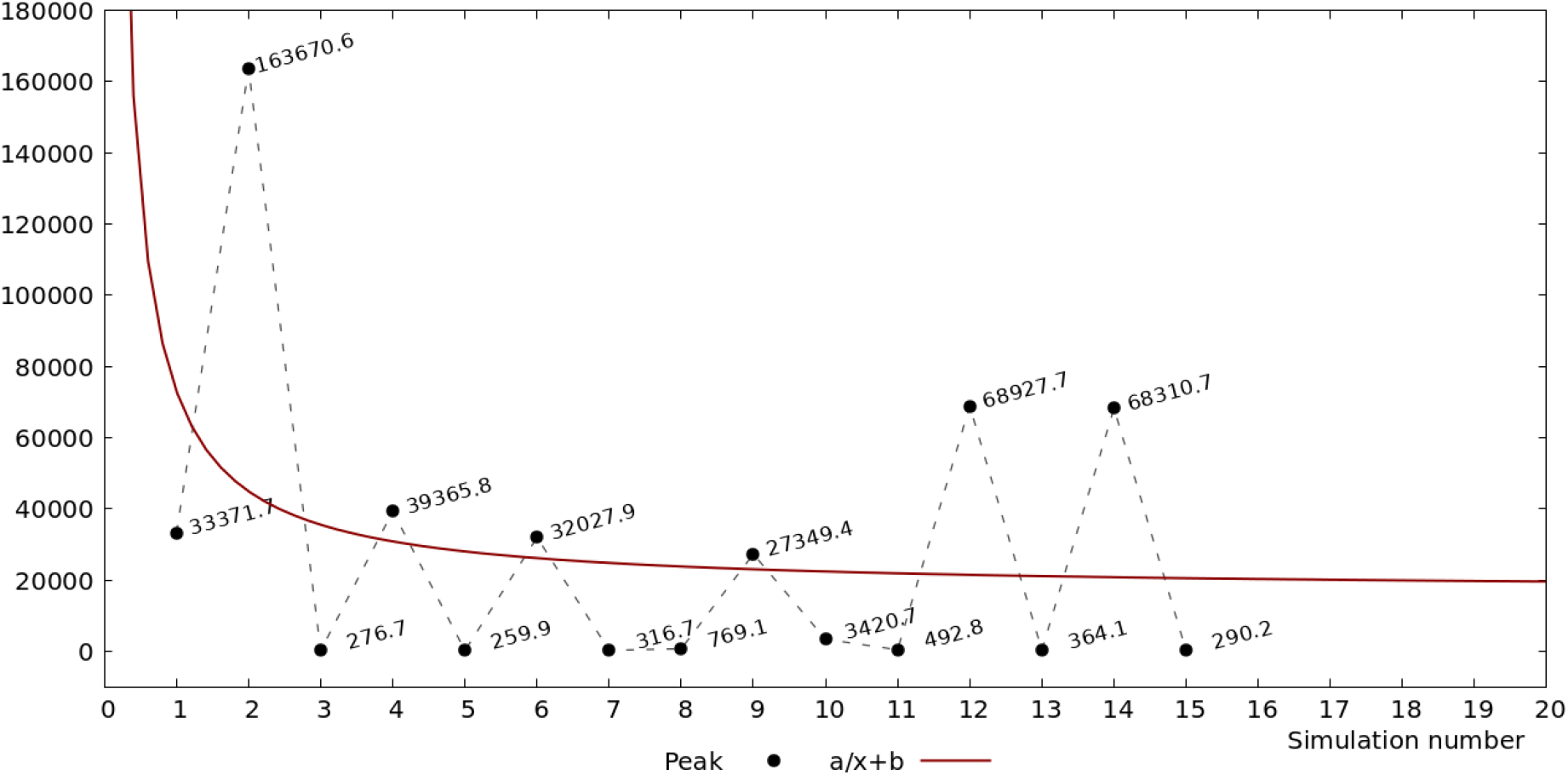
Maximum values (peaks) obtained in numerical simulations in each 14-day period. **Source:** The authors.

Complementarily, in Figure **??** we present a scenario of possible infection peaks that could happen. For each of the 15 simulations we estimated the maximum number of infected individuals. The black dots are the peak values of each of our simulations, and the red curve is a proposed least squares fit (BURDEN;FAIRES, 2011) with *a* = 62057 and *b* = 13328 parameters. The red curve shows, on average, how the city is experiencing the Covid-19 epidemic.

## Discussion

Data from the Department of Health of Londrina4 show how the behavior of the disease can abruptly changing in short periods. These changes are due to a series of socio-political factors, whose mathematical quantification is still a challenge.

However, by the way we modeled and numerically solved the COVID-19 problem in Londrina, these changes can be adequately mapped, for periods of 14 days, through a dynamic parameter *β*, via mathematical modeling. This strategy allows us to visualize future scenarios, described not only by punctual results, but by a historical series of the infection’s behavior, and thus support decision-making.

One of the elements that signal the dynamics of these changes is the reproduction number of the disease *R*_*t*_. In this way, we emphasize the importance of evaluating the variation of *R*_*t*_ over the days as a means of supporting the effectiveness of measures adopted to contain the disease in Londrina.

Another complementary element that guides the dynamics of abrupt changes are the peaks of the numerical simulations for the numbers of infected individuals. This element tells us the potential capacity of infected people that could come to exist, and consequently, how much this could impact the existing health system in the city. Finally, as the epidemic is in progress, researchers at the Laboratory of Simulation and Numerical Analysis (LabSAN), from the Mathematics department at the State University of Londrina, are maintaining and disseminating the results and potential consequences on the spread of COVID-19 in the city Londrina over time.The results can be visualized in <http://www.uel.br/laboratorios/labsan/covid.html>.

## Data Availability

Declaration of data availability: All data mentioned in the manuscript and the links below are available.

## Acknowledgments

This study was financed in part by the Coordenação de Aperfeiçoamento de Pessoal de Nível Superior - Brasil (CAPES) - Finance Code 001.

## Footnotes

1 <https://saude.londrina.pr.gov.br/index.php/dados-epidemiologicos/boletim-informativo.html>

2 First day on which the number of recovered (removed) was made available by the Daily Bulletin.

3 <https://saude.londrina.pr.gov.br/index.php/dados-epidemiologicos/boletim-informativo.html>

4 <https://saude.londrina.pr.gov.br/index.php/dados-epidemiologicos/boletim-informativo.html>

